## Supplementary material for "All-cause and cause-specific mortality during and following incarceration in Brazil: a retrospective cohort study": S1 Appendix

**Fuzzy matching within SIGO incarceration database**

Individuals in the SIGO database were identified by name, mother’s name, and RGI number (an internal identification number). However, mother’s name was missing for the majority of entries before 2015, and RGI number was missing in 144,813 entries. Thus, to identify unique individuals, we compared name and RGI number, using mother’s name (when present) to check for consistency.

When a single RGI number was associated with different names and/or different mother’s names, or when the same name was associated with different or missing RGI numbers, we performed fuzzy string matching on names and mother’s names using the *fuzzywuzzy* package in Python **(Fig. S1)**. We computed a string similarity score adapted from the WRatio (weighted ratio) function as follows. First, we computed a standard ratio based on Levenshtein distance. Then, if one name was at least 1.2 times longer than the other (for example, due to a middle name), we computed a substring similarity ratio and scaled it by 0.95 (0.9 for mother’s names); if one name was more than twice as long as the other, we scaled the similarity ratio by 0.85. We then took the larger value of the standard ratio or scaled substring ratio as the string similarity score.

We determined a string similarity threshold for names (78% similarity) by examining a subset of entries with the same RGI number but different names that we manually confirmed to correspond either to the same individual (“consistent names”) or to different individuals. We determined a separate threshold for mother’s names (85% similarity) by manually examining a subset of entries with consistent names but different mother’s names.

Using these thresholds, we compared all entries with the same RGI number but different names, and all entries with the same name but different RGI numbers (Fig. S1). All entries determined to correspond to the same individual were evaluated against an additional, more stringent threshold (90% similarity). Entries that fell below that threshold for name or mother’s name were manually examined and were either verified to correspond to the same individual or were corrected to be different individuals. After applying these rules, there were 143,551 unique individuals with at least one recorded movement in SIGO between 2005 and 2018.

**Inferring sex of individuals in SIGO**

To infer the sex of individuals in SIGO for sex-stratified analyses, we compiled a list of individuals who had at least one incarceration in a female prison. Due to changes in some facilities housing different sexes over time, we considered individuals to be male if they had at least one incarceration in a male prison during the study period. We then manually evaluated and corrected sex classifications based on name. Data were insufficient to determine whether individuals were transgender or gender non-conforming.

**Converting SIGO movement data to interval data**

We summarized incarceration history for the 143,551 individuals by converting movement data into interval data by facility type. For example, if an individual were transferred to facility (type A) on 01/01/2010, transferred to another facility (type A) on 01/15/2010 and released on 02/01/2010, we considered that individual to be in facility type A from 01/01/2010 to 02/01/2010 and in the community from 02/01/2010 until another movement, death, or the end of the study period. Movement data were incomplete; therefore, we considered a movement as a release if it was not clearly recorded as an “exit” (defined below) and if the destination of the movement was blank or to a location different than the origin location of that individual’s next record in SIGO. Clearly recorded exits were those that were annotated as alvara de soltura, evasão, fuga, óbito, permissão de saída, habeas corpus, flabeus corpus, saída temporária, or liberação condicional.

**Truncation of intervals in facilities with underreported releases**

We observed in the interval data that the population in police stations, semi-open prisons, and youth detention facilities appeared to be continually increasing in a manner that was inconsistent with DEPEN’s reported size of detained populations in these facilities. We attributed this discrepancy to SIGO’s under-documentation of releases from these facility types. To account for this, we computed the 0.9 quantile of the distribution of time spent in each of these facility types among intervals with clearly recorded exits (109 days, 320 days, and 459 days in men and boys in police stations, semi-open prisons, and youth detention facilities, respectively; 224 days and 434 days for women in police stations and semi-open prisons, respectively). Then, for intervals in these facility types where release was not clearly documented, we truncated intervals at the 0.9 quantile timepoint, after which individuals were marked as being in the outside community. We excluded intervals with these artificial “releases” from post-release hazard and survival analyses.

**Fuzzy matching between SIGO and SIM**

For fuzzy matching of names and mother’s names between SIGO and SIM, we computed similarity scores as described above and compiled all putative matches with >= 85% similarity between names and >= 80% similarity between mother’s names **(Fig. S2)**. We filtered putative matches based on date, such that matches were removed if an individual had a record in SIGO more than 30 days after the death date of their supposed match in SIM, unless mother’s names were consistent and the combined similarity score was greater than or equal to 90%. We then ranked putative matches for each death in SIM by the difference between the date of death in SIM and the date of an individual’s last record in SIGO. Next, we selected the best match based on similarity score for name and mother’s name. We then filtered matches using string similarity thresholds optimized for positive predictive value, assuming a 4.5% prevalence of incarceration/incarceration history based on our cumulative person-time data. For sensitivity, we considered matches in which deaths were documented in both SIM and SIGO to be our “gold standard” of true positives. While 461 deaths were documented in SIGO between 2005 and 2018, only 402 had putative “best” matches in SIM, of which 388 were manually verified to be true matches based on name, mother’s name, and date of death. Therefore, the maximum sensitivity possible was 84.2%. For specificity, we assumed that children under 12 would not be incarcerated and thereby considered matches to be false positives if age at death was under 12 years. We determined two optimal thresholds for similarity score: one for when mother’s name was present in SIGO and the threshold would be applied to the average similarity score for name and mother’s name (91.5%), and the other for when mother’s name was absent in SIGO and the similarity threshold was applied to the similarity score between names only (99%). Our final thresholds yielded a sensitivity of 76.6%, specificity of 99.8%, and positive predictive value of 94.7%. We then excluded deaths for which mother’s name was missing and there were multiple perfect matches by name only (N=771). As this led to a substantial reduction in the total number of identified matches, we examined the sensitivity of our findings to the inclusion/exclusion of these uncertain matches (see Sensitivity Analysis). Next, we manually removed 21 matches where an individual would have been less than 12 years old at their first incarceration. We additionally inspected matches that had a similarity score under 90% for name but had passed filtering due to high similarity of mother’s name; we removed 34 of these “sibling” matches. We further removed 4 matches which had no age information in SIM but had “neonatal conditions” as the recorded cause of death. We added back 70 matches corresponding to deaths documented in SIGO that had been manually verified but had not met the similarity score threshold or had been filtered out from the steps above. We also added 51 deaths documented in SIGO that had no match in SIM; these deaths were excluded from age- and cause-based analyses. We limited the study period to 2009-2018 and excluded deaths of individuals under 14 or over 95 years of age, resulting in 5,859 total matches between SIGO and SIM that were excluded to get deaths of non-incarcerated individuals (Fig. 1). We further excluded 81 deaths in uncertain locations, 2638 deaths following release from locations other than prison, 11 deaths of individuals under 18 or over 95 years old (except those in youth detention), and 2 deaths of individuals over 19 in youth detention. This resulted in 3127 deaths during and after incarceration for downstream analysis.

**Cause of death categories**

The Global Burden of Disease Study classified many ICD-10 codes as “garbage codes” if they were not considered underlying causes of death or were incompletely reported (i.e. “neoplasms, unspecified site”); deaths with these codes were redistributed by age, sex, location, and year to the most likely causes of death. However, given the absence of reliable data on mortality during and following incarceration, we could not perform redistribution and instead sought to include as many causes as possible, even if incompletely reported. We therefore used the following classifications for “garbage codes.” Hanging, strangulation and suffocation, undetermined intent was classified as interpersonal violence. Deaths were classified as cardiovascular disease if they were reported as ischemic heart disease, rheumatic heart disease, cerebrovascular disease, pulmonary embolism, essential (primary) hypertension, heart failure, atherosclerosis, cardiac arrest, arterial embolism and thrombosis, disseminated intravascular coagulation, atrioventricular and left bundle-branch block, other circulatory diseases, other cardiac arrhythmias, or complications and ill-defined descriptions of heart disease. Renal failure was classified under skin/genitourinary/musculoskeletal diseases. HIV/AIDS resulting in malignant neoplasms was classified under neoplasms. Other disorders of brain were classified under mental/behavioral, neurological, and sense organ disorders. Other diseases of digestive system were classified under other digestive diseases.

We assigned intermediate cause categories to broad cause categories as follows. Deaths classified under interpersonal violence, self-inflicted injuries, or transport injuries were directly attributed to violence, suicide, or transport injuries, respectively. Deaths were attributed to communicable diseases if they were classified as HIV/AIDS, STDs excluding HIV, respiratory infections, tuberculosis, hepatitis, meningitis and encephalitis, parasitic and vector-borne diseases, intestinal infectious diseases, malaria, sepsis, rheumatic heart disease, selected vaccine preventable diseases, or other infectious diseases. Deaths were attributed to non-communicable diseases if they were classified under cardiovascular disease; neoplasms; liver disease; mental/behavioral, neurological, and sense organ disorders; respiratory diseases; skin/genitourinary/musculoskeletal diseases; nutritional deficiencies; endocrine, nutritional, blood and immune disorders; congenital anomalies; or other digestive diseases.

**Estimation of age structure for incarcerated population and population without recent incarceration**

We started with age structure data reported in wide bins that were not directly comparable between the incarcerated population and the population without recent incarceration. To enable more precise calculation and comparison of age-specific and age-standardized rates between populations, we fit truncated negative binomial distributions to the binned data using maximum likelihood estimation, implemented through the *fitdistcens* function from the R package *fitdistrplus* and adapted for truncated distributions using the *truncdist* package. We estimated starting mean and dispersion values using the *revengc* package. We compared the fitted distributions to the binned data and re-ran the estimation procedure with adjusted weights for certain age groups in order to improve the fit. For instance, we observed that the initial distribution of the incarcerated population underestimated the proportion of those in the 25-29 age group, which was also the most populous according to DEPEN in most years; we therefore set that age group to have twice the weight of other age groups in the estimation procedure. To further improve the consistency of our estimates with reported age structure, we adjusted the estimated counts to correspond proportionally to the reported binned data. For example, if the age structure was reported such that the 25-29 age group comprised 25% of the incarcerated population in 2012, we scaled our modeled counts for each year in the 25-29 age group to sum to 25% of the total incarcerated person-time in 2012. This way, we utilized the shape within each age bin, as derived from the fitted distribution, while retaining the reported proportions within each age group. For reported age bins that were extremely wide (i.e. age 46-60), this enabled more granular, accurate comparison of mortality rates across populations with starkly different age compositions.

**Sensitivity analysis**

We examined the sensitivity of all-cause and cause-specific mortality rates to the inclusion or exclusion of matches for which mother’s name was missing and there were multiple perfect matches by name. Out of the multiple perfect matches, we selected the one with the minimum date difference between death date and an individual’s last record in SIGO. This led to 764 additional deaths from 2009-2018 of individuals between the ages of 14 and 95 (31 in closed prisons, 3 in semi-open prisons, 13 in police stations, 682 post-release, and 35 in other/unknown locations). We assessed the effect of including these uncertain matches on all-cause and cause-specific mortality rate ratios for men and women. Inclusion of these uncertain matches led to higher rate ratios for deaths from non-communicable and communicable diseases, particularly among older age groups **(Fig. S12)**, but did not substantially change our conclusions.
