## Supplementary material for "All-cause and cause-specific mortality during and following incarceration in Brazil: a retrospective cohort study": Table S1

| Population | | Facility type/location | Analyses (figure) | Reference population |
|---|---|---|---|---|
| Incarcerated | Men | Closed prisons | Age-specific (Fig. S6) and age-standardized (Fig. 3) rates; instantaneous hazard (Fig. S8) and binned age-standardized rates (Fig. S9) | Non-incarcerated men |
| | | Semi-open prisons | Age-specific (Fig. S6) and age-standardized (Fig. S7) rates; instantaneous hazard (Fig. S8) and binned age-standardized rates (Fig. S9) | |
| | | Police stations | Age-specific (Fig. S6) and age-standardized (Fig. S7) rates; instantaneous hazard (Fig. S8) and binned age-standardized rates (Fig. S9) | |
| | Boys | Youth detention | Crude (age-specific) rates (Fig. S7) | Non-incarcerated boys (age 14-19) |
| | Women | All facility types (closed prisons, semi-open prisons, police stations) | Age-standardized rates (Fig. 3) | Non-incarcerated women |
| Post-release | Men | Released from prison (closed or semi-open) | Age-specific (Fig. S6) and age-standardized (Fig. 4) rates; instantaneous hazard (Fig. 5); Kaplan-Meier survival, cause-of-death, and time-to-death, stratified by total time incarcerated (Fig. S11) | Non-incarcerated men |
| | Women | Released from prison (closed or semi-open) | Age-specific (Fig. S10A) and age-standardized (Fig. 4) rates; instantaneous hazard (Fig. S10B) | Non-incarcerated women |
