## Supplementary material for "All-cause and cause-specific mortality during and following incarceration in Brazil: a retrospective cohort study": Table S2

| | Incarcerated Men and Boys | Formerly Incarcerated Men and Boys | Incarcerated Women | Formerly Incarcerated Women | All groups |
|---|---|---|---|---|---|
| **All causes** | 672 | 2236 | 33 | 186 | 3127 |
| **Interpersonal violence** | 226 (33.6%) # | 848 (37.9%) # | 2 (6.1%) # | 31 (16.7%) # | 1107 (35.4%) # |
| **Cardiovascular disease** | 79 (11.8%) # | 311 (13.9%) # | 7 (21.2%) # | 29 (15.6%) # | 426 (13.6%) # |
| **Transport injuries** | 28 (4.2%) | 205 (9.2%) # | 0 | 7 (3.8%) | 240 (7.7%) # |
| **Neoplasms** | 24 (3.6%) | 139 (6.2%) # | 3 (9.1%) # | 34 (18.3%) # | 200 (6.4%) # |
| **Suicide** | 55 (8.2%) # | 90 (4.0%) # | 3 (9.1%) # | 7 (3.8%) | 155 (5.0%) # |
| **Respiratory infections (including TB)** | 50 (7.4%) # | 78 (3.5%) | 0 | 8 (4.3%) # | 136 (4.3%) |
| **HIV/AIDS** | 30 (4.5%) # | 79 (3.5%) | 7 (21.2%) # | 11 (5.9%) # | 127 (4.1%) |
| **Other/unknown causes** | 180 (26.8%) | 486 (21.7%) | 11 (33.3%) | 59 (31.7%) | 736 (23.5%) |
