## Supplementary figures and images for "All-cause and cause-specific mortality during and following incarceration in Brazil: a retrospective cohort study"

### Fig S1

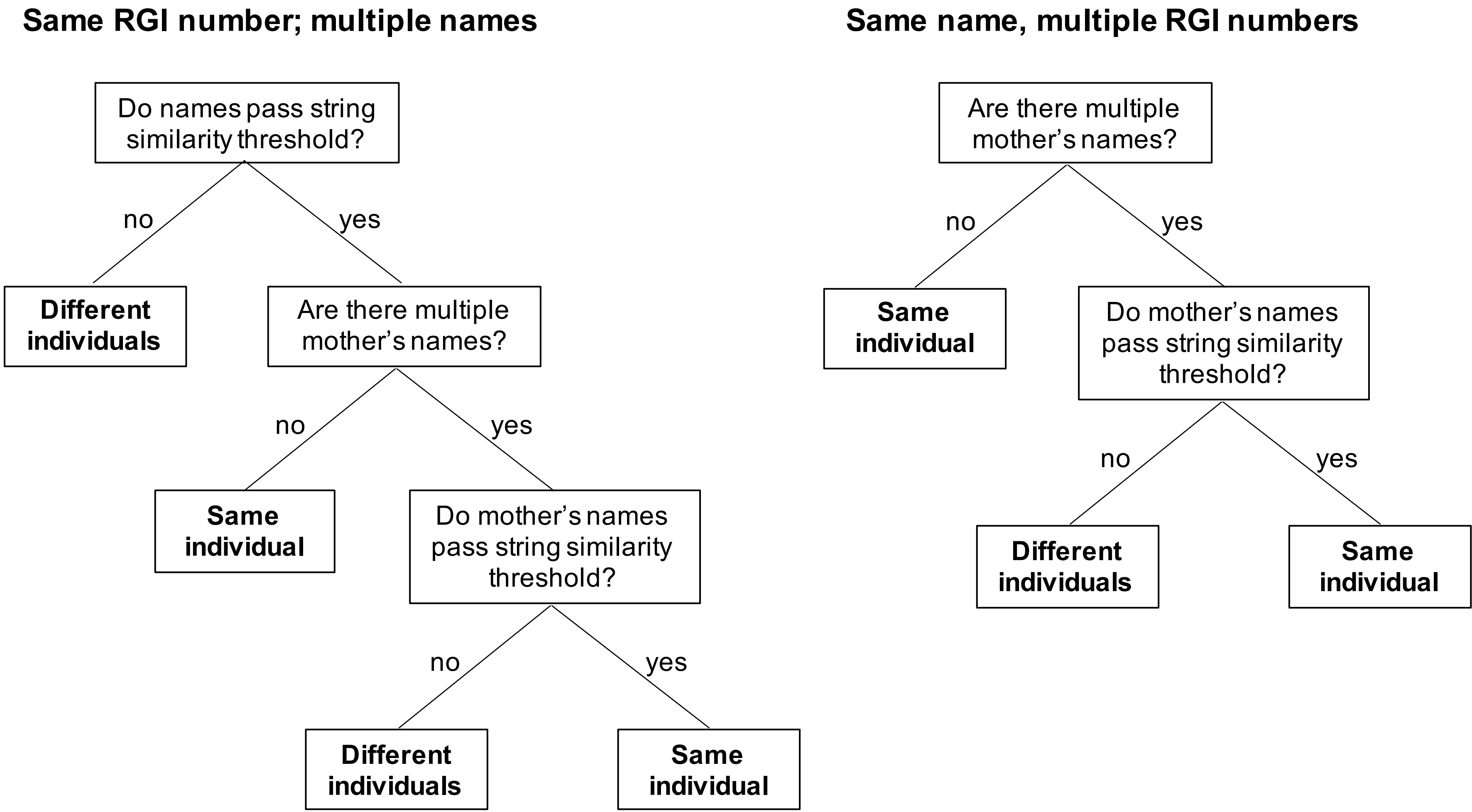

### Fig S2

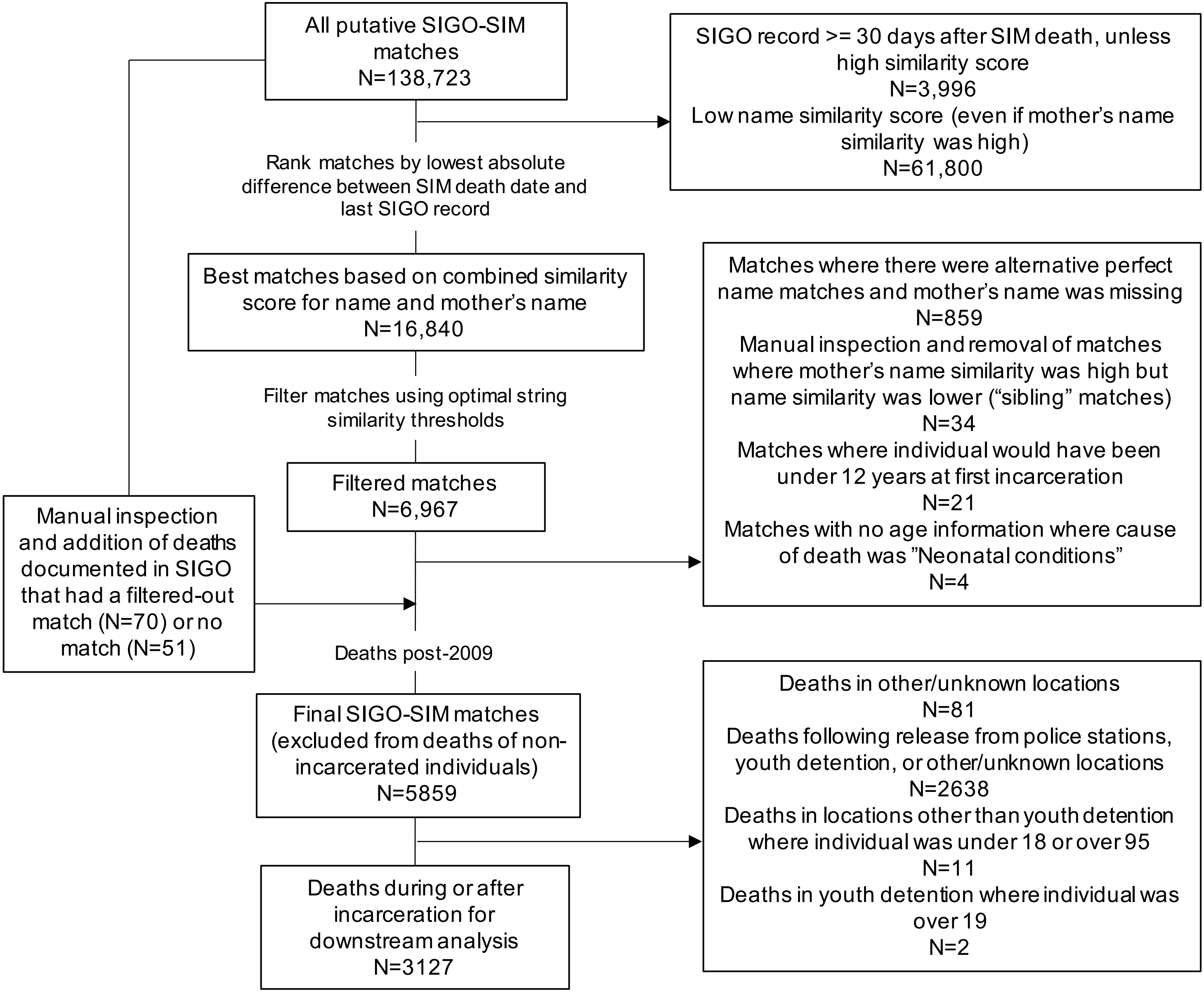

### Fig S3

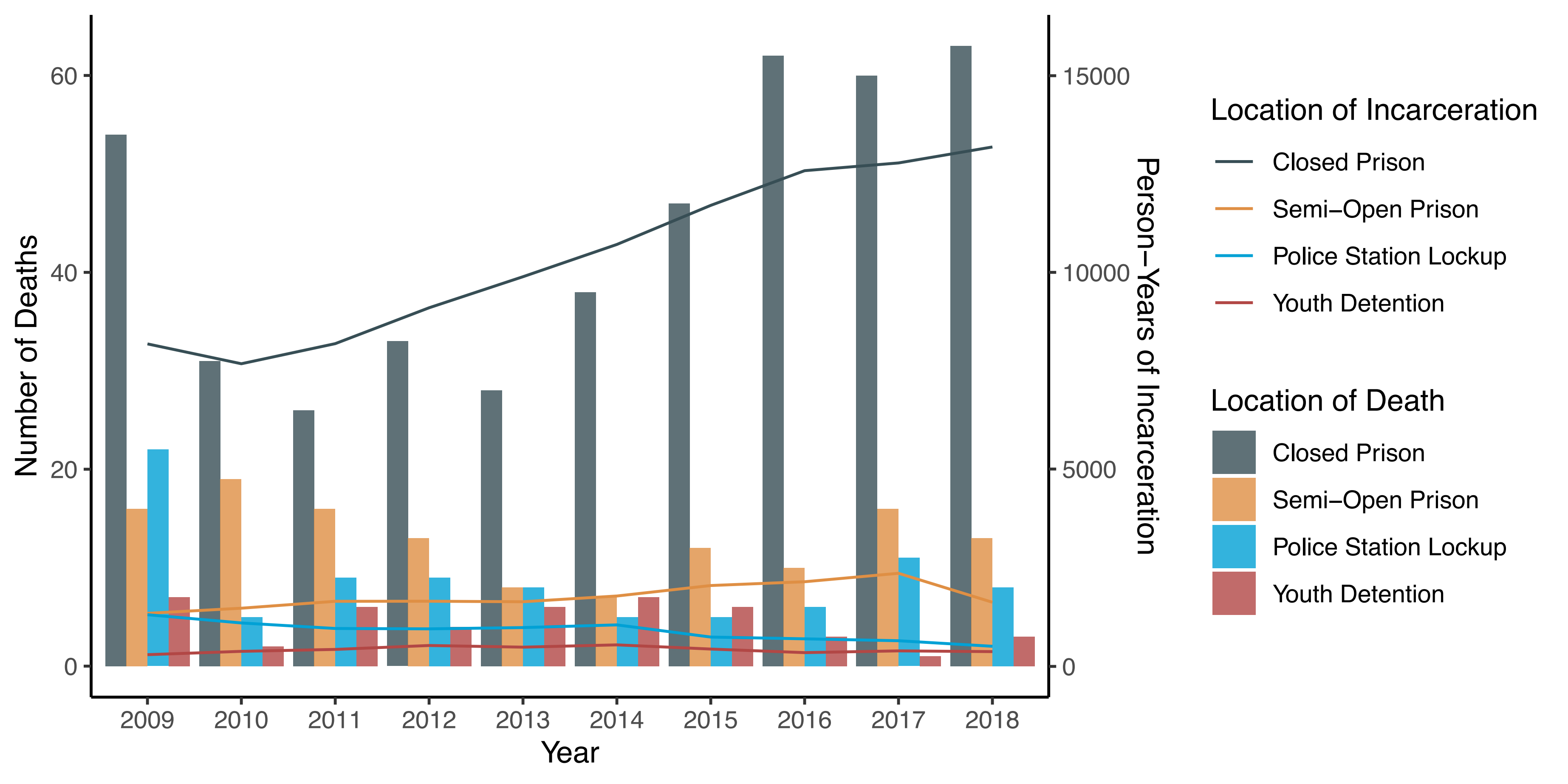

### Fig S4

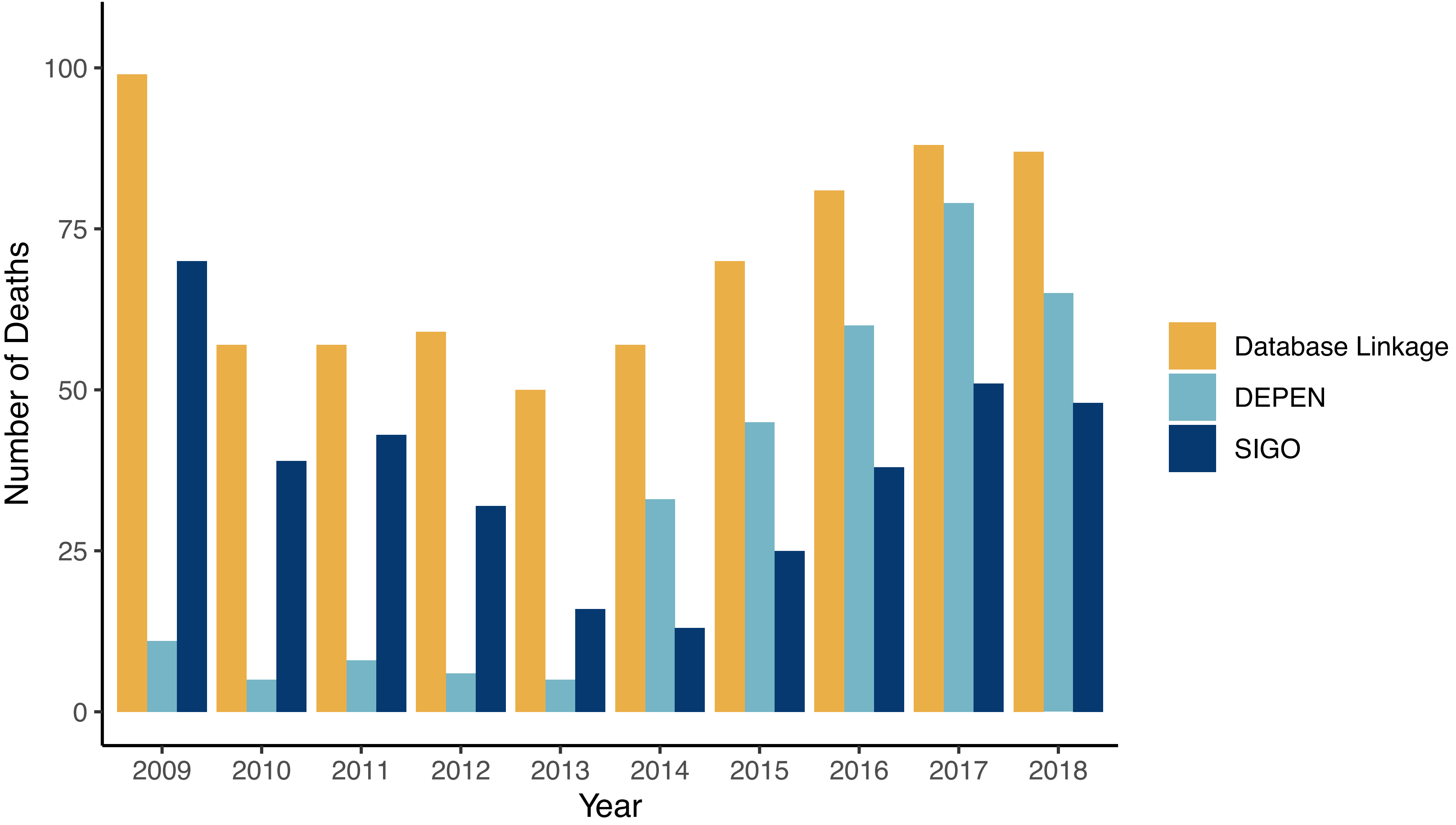

### Fig S5

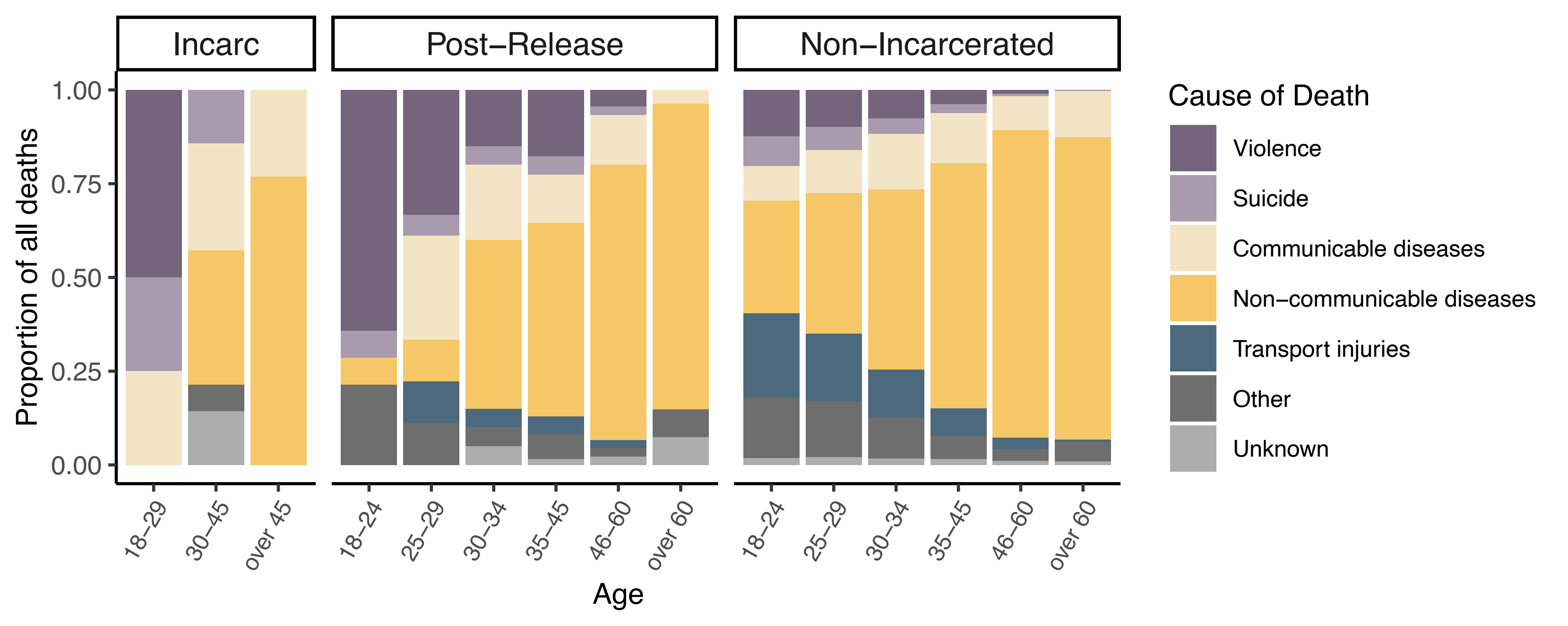

### Fig S8

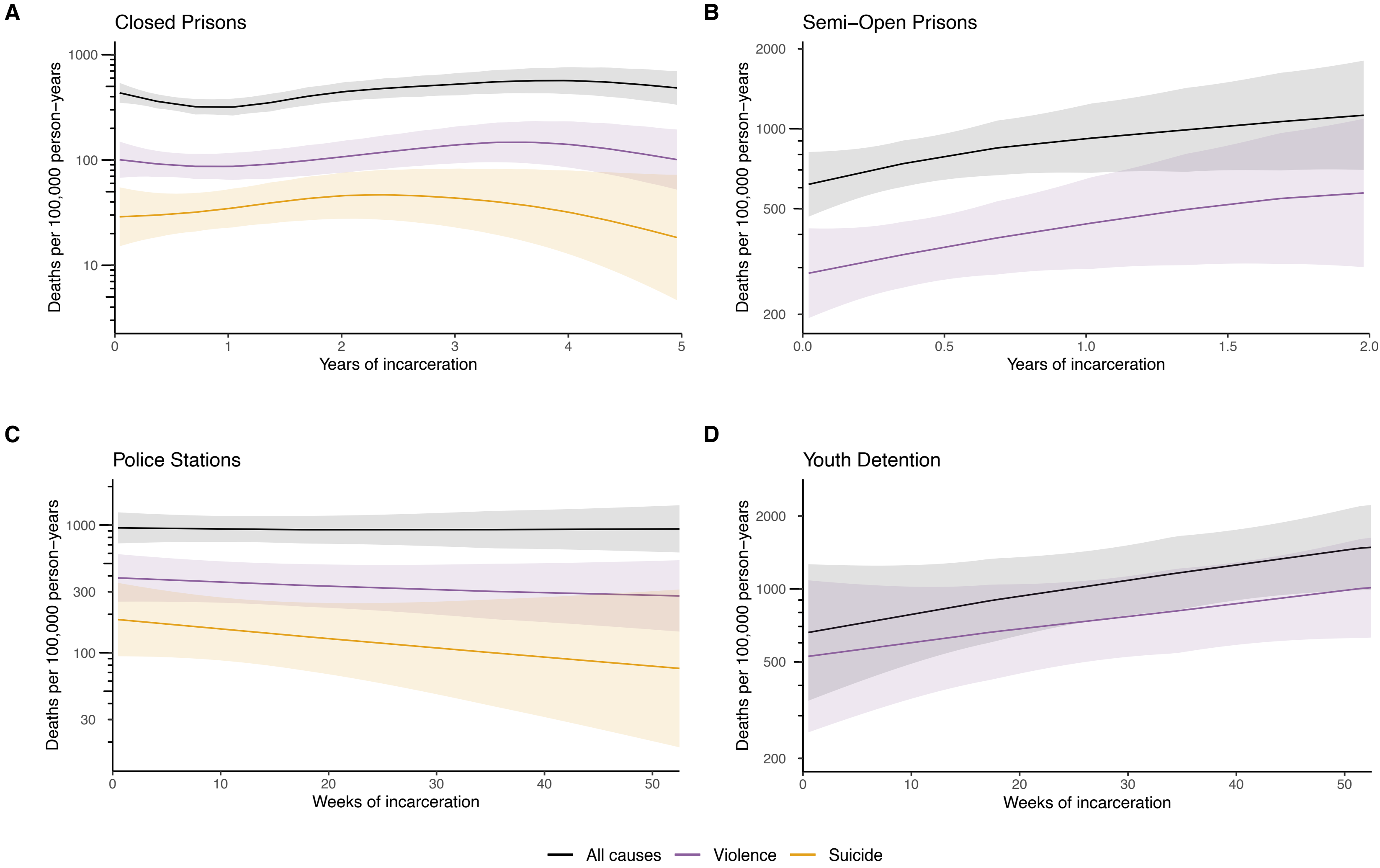

### Fig S9

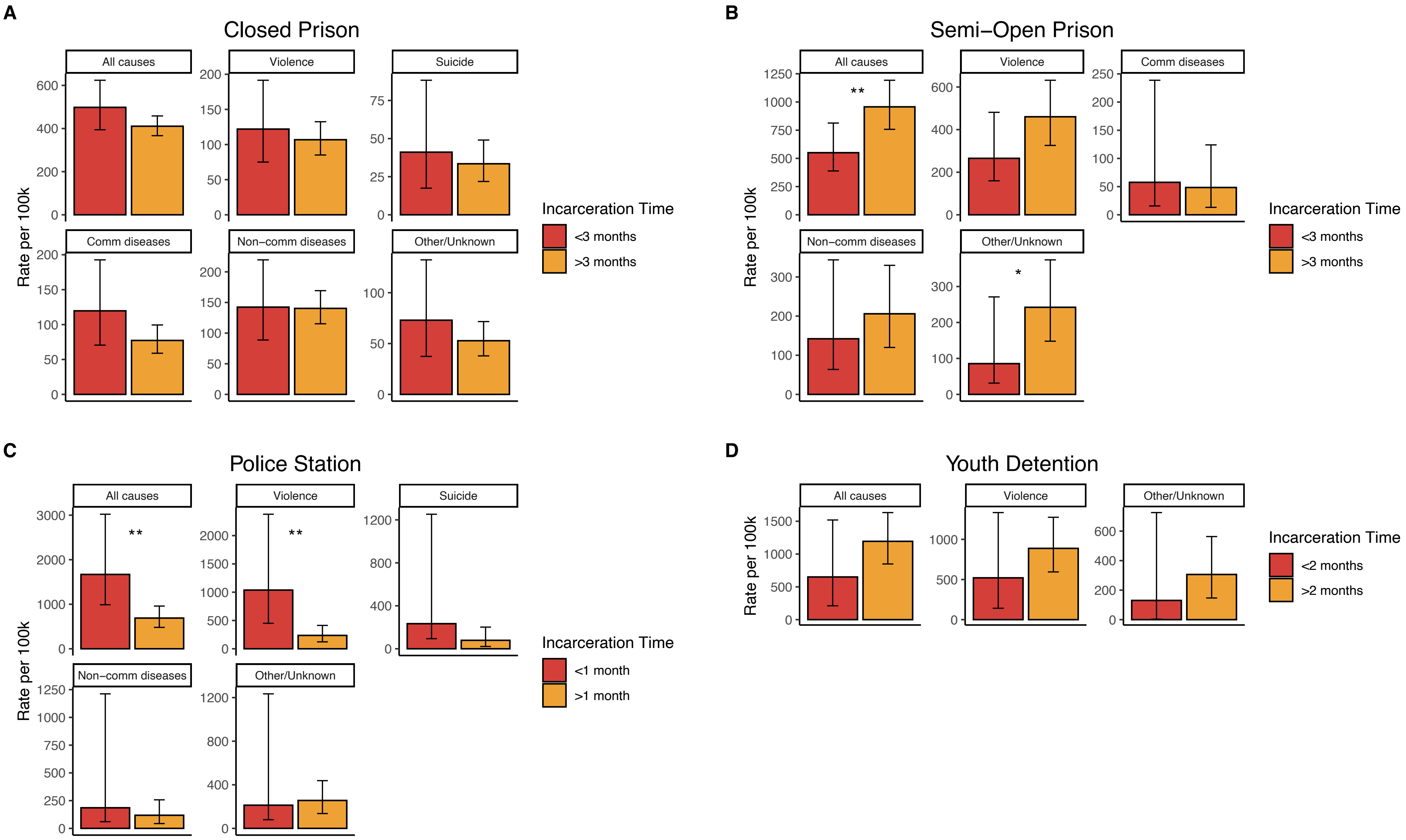

### Fig S10

**A**

| Cause | Age | N | Rate per 100K | IRR (95% CI) |
|---|---|---|---|---|
| All causes | 18–24 | 14 | 225.7 | 3.4 (1.8–5.7) |
| | 25–29 | 18 | 202.8 | 2.6 (1.5–4.1) |
| | 30–34 | 20 | 215.7 | 2 (1.2–3.2) |
| | 35–39 | 27 | 355.4 | 2.5 (1.6–3.7) |
| | 40–44 | 29 | 581.2 | 3 (2–4.3) |
| | 45–50 | 17 | 533.8 | 1.8 (1–2.8) |
| | 50–60 | 25 | 733.6 | 1.5 (0.9–2.2) |
| | 60–70 | 23 | 3125 | 2.9 (1.8–4.3) |

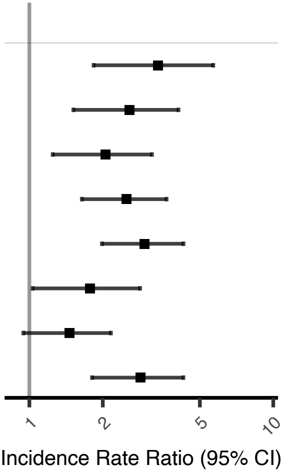


**B**

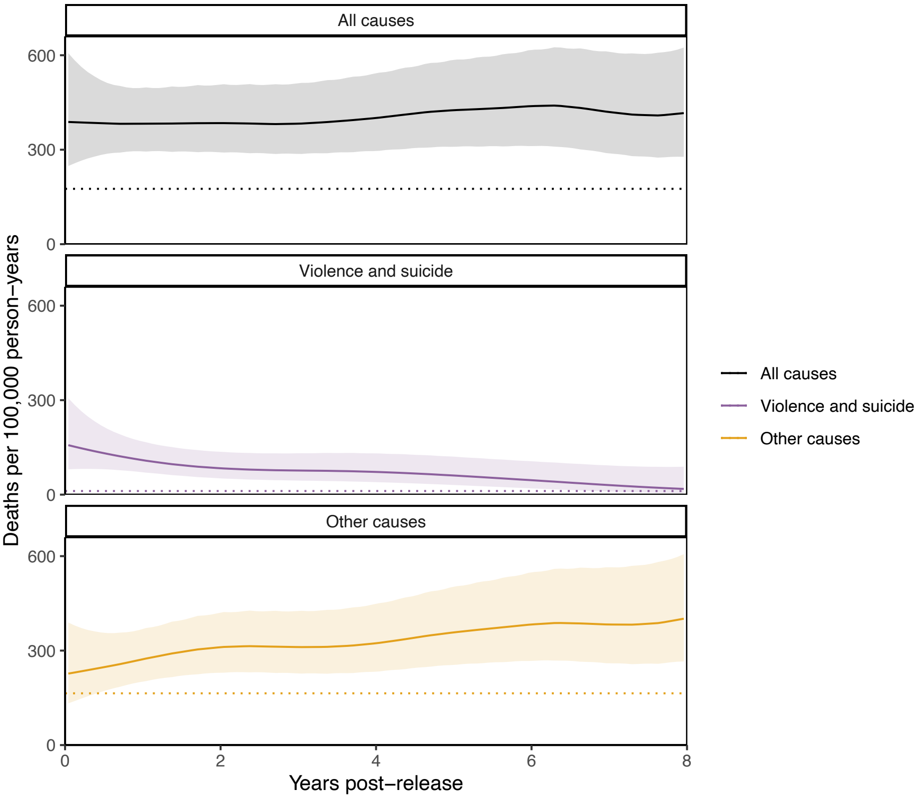

### Fig S11

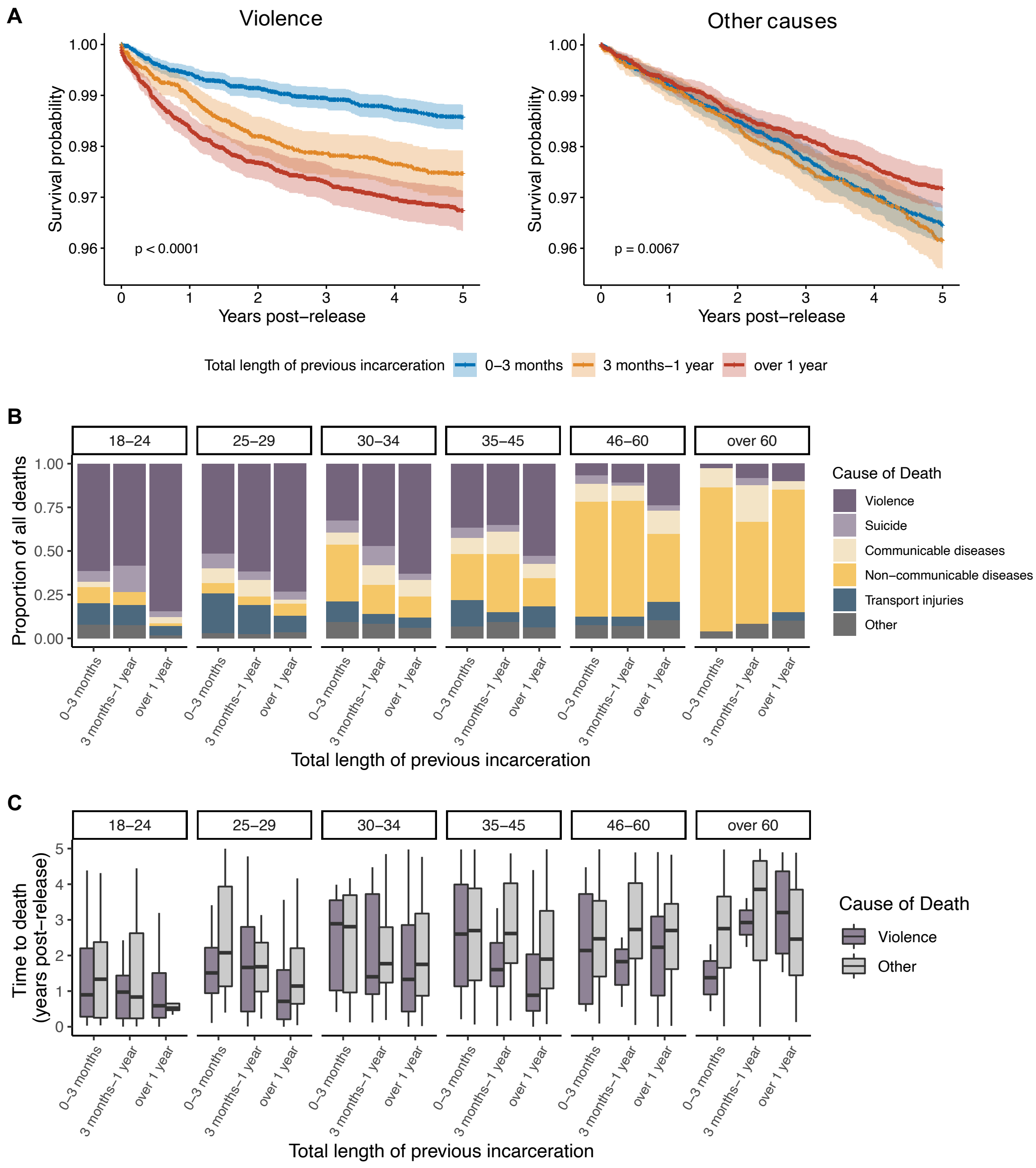

### Fig S12

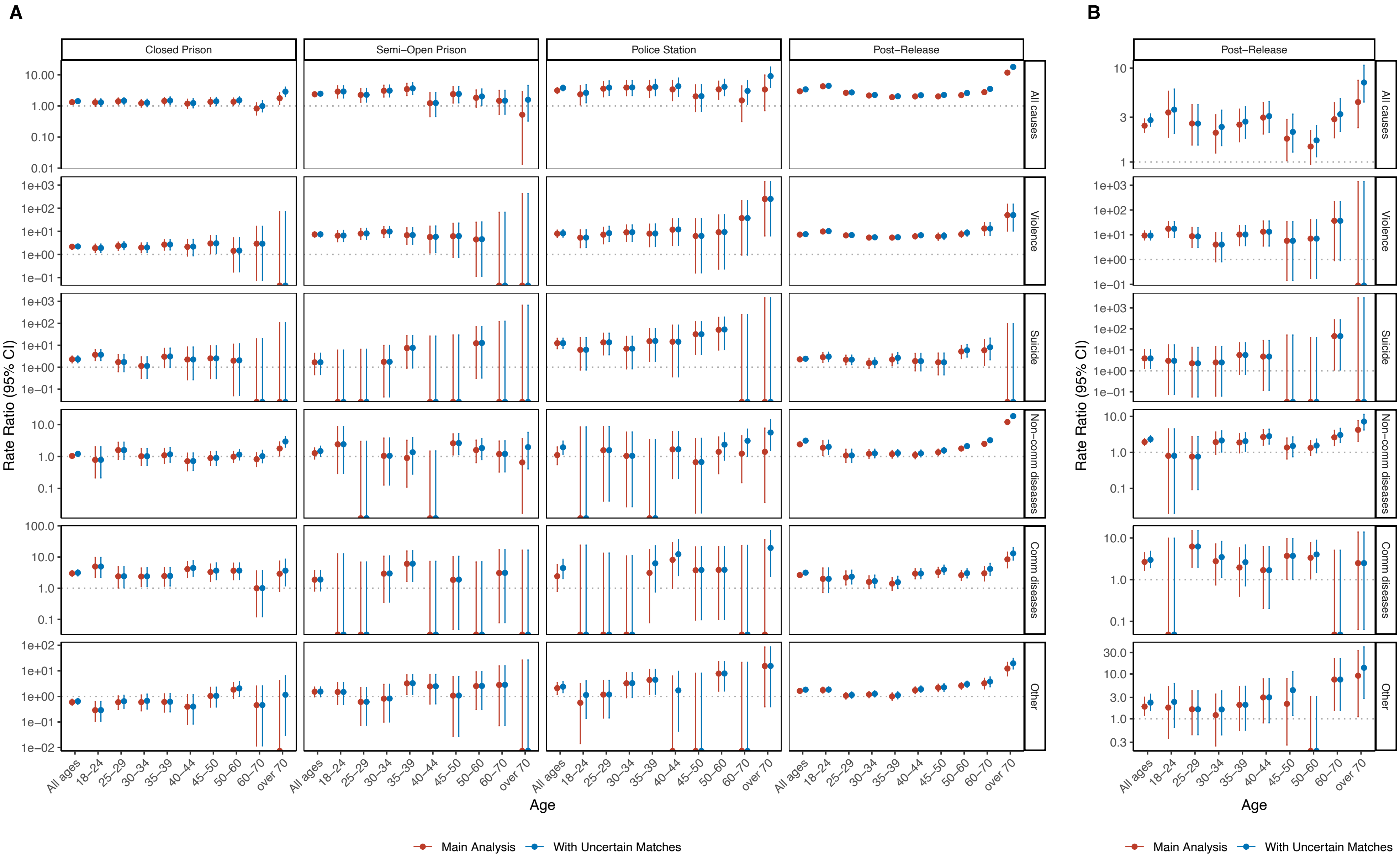

### Fig S13

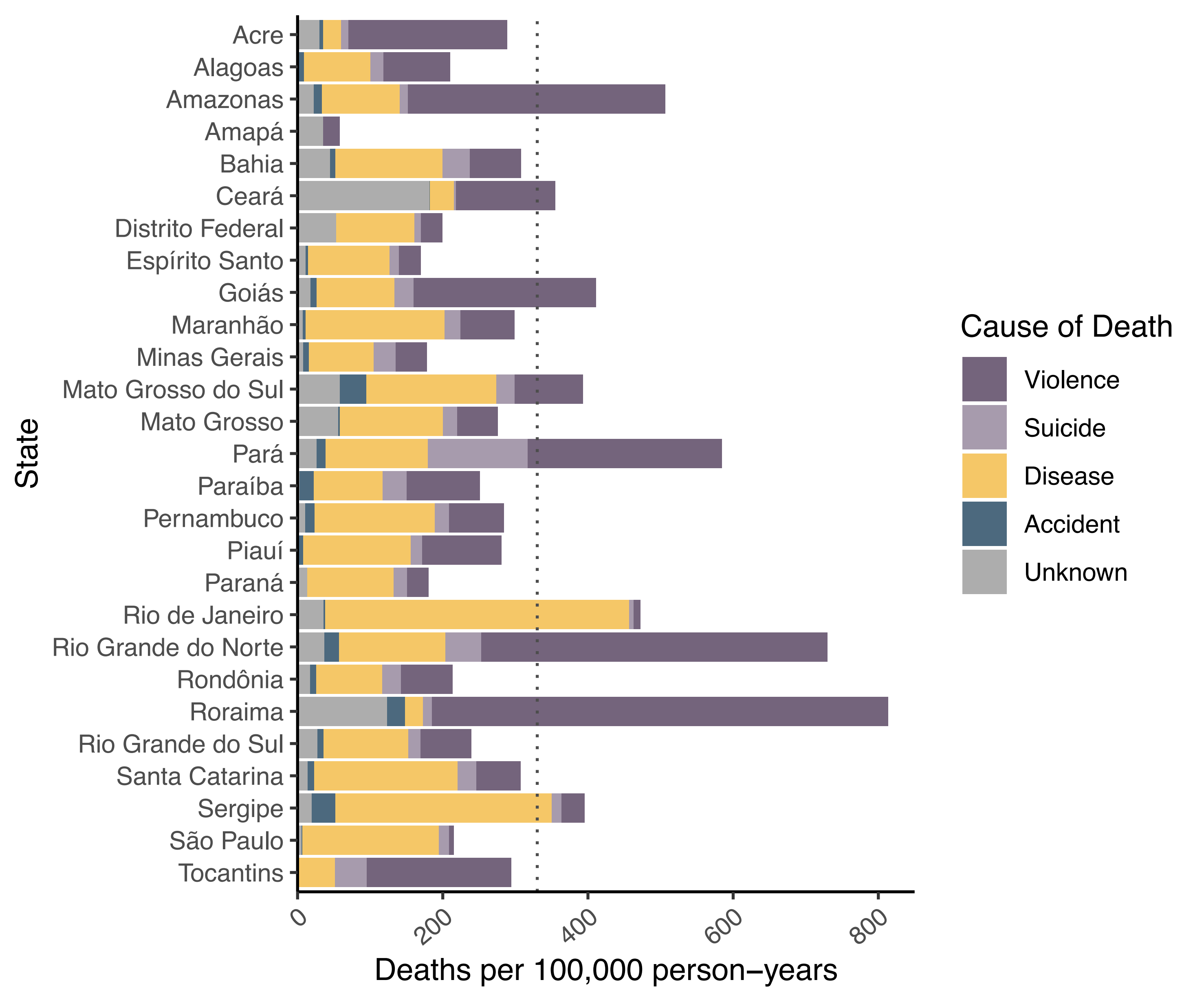
