## Supplementary material for "All-cause and cause-specific mortality during and following incarceration in Brazil: a retrospective cohort study": Fig S6

### Closed Prison

| Cause | Age | N | Rate per 100K | IRR (95% CI) |
|---|---|---|---|---|
| All causes | 18–24 | 58 | 263.3 | 1.3 (1–1.7) |
| | 25–29 | 65 | 281.3 | 1.4 (1.1–1.8) |
| | 30–34 | 54 | 287 | 1.2 (0.9–1.6) |
| | 35–39 | 54 | 427.5 | 1.4 (1.1–1.9) |
| | 40–44 | 36 | 475.6 | 1.2 (0.8–1.6) |
| | 45–50 | 42 | 764.1 | 1.4 (1–1.8) |
| | 50–60 | 51 | 1300.4 | 1.4 (1–1.8) |
| | 60–70 | 21 | 1734.1 | 0.8 (0.5–1.3) |

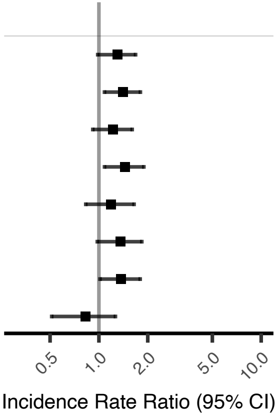


### Semi–Open Prison

| N | Rate per 100K | IRR (95% CI) |
|---|---|---|
| 21 | 587.7 | 2.9 (1.8–4.4) |
| 17 | 453.2 | 2.3 (1.3–3.6) |
| 22 | 721.3 | 3.1 (1.9–4.7) |
| 21 | 1029.4 | 3.5 (2.2–5.3) |
| 6 | 491.4 | 1.2 (0.4–2.7) |
| 12 | 1351.4 | 2.4 (1.2–4.2) |
| 11 | 1740.5 | 1.8 (0.9–3.3) |
| 6 | 3061.2 | 1.5 (0.5–3.2) |

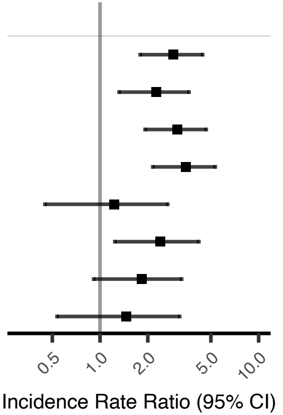


### Police Station

| Cause | Age | N | Rate per 100K | IRR (95% CI) |
|---|---|---|---|---|
| All causes | 18–24 | 9 | 479 | 2.4 (1.1–4.5) |
| | 25–29 | 14 | 719.8 | 3.6 (2–6.1) |
| | 30–34 | 14 | 915.6 | 3.9 (2.1–6.6) |
| | 35–39 | 11 | 1103.3 | 3.7 (1.9–6.7) |
| | 40–44 | 8 | 1362.9 | 3.4 (1.5–6.7) |
| | 45–50 | 5 | 1144.2 | 2 (0.7–4.7) |
| | 50–60 | 10 | 3246.8 | 3.4 (1.6–6.3) |
| | 60–70 | 3 | 3157.9 | 1.5 (0.3–4.4) |

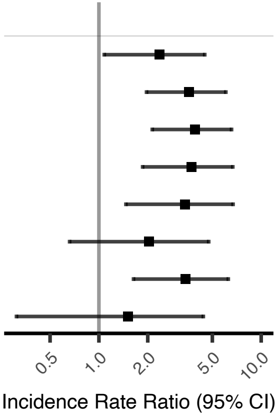


### Post–Release

| N | Rate per 100K | IRR (95% CI) |
|---|---|---|
| 317 | 867.7 | 4.3 (3.8–4.8) |
| 300 | 528.6 | 2.6 (2.3–3) |
| 299 | 507.7 | 2.2 (1.9–2.4) |
| 249 | 565.4 | 1.9 (1.7–2.2) |
| 218 | 811.6 | 2 (1.8–2.3) |
| 189 | 1140.8 | 2 (1.7–2.3) |
| 318 | 2116.3 | 2.2 (2–2.5) |
| 187 | 5820.1 | 2.8 (2.4–3.2) |

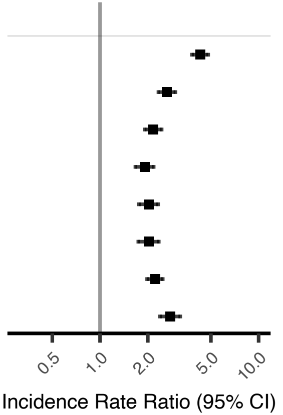
