## Supplementary material for "All-cause and cause-specific mortality during and following incarceration in Brazil: a retrospective cohort study": Fig S7

### Semi–Open Prison

| Cause | N | Rate per 100K | IRR (95% CI) |
|---|---|---|---|
| All causes | 117 | 761.5 | 2.4 (2–2.8) |
| Violence | 57 | 370.5 | 7.3 (5.5–9.5) |
| Suicide | 4 | 26.1 | 1.7 (0.5–4.3) |
| Non–communicable diseases | 26 | 169.5 | 1.3 (0.8–1.9) |
| Communicable diseases | 8 | 52.2 | 1.9 (0.8–3.7) |
| Other | 22 | 143.3 | 1.5 (1–2.3) |

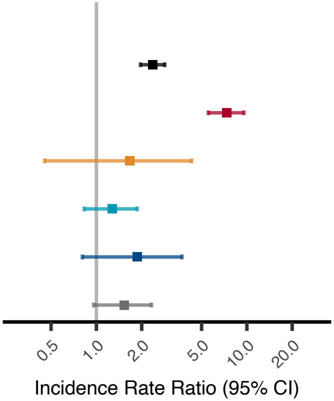


### Police Station Lockup

| Cause | N | Rate per 100K | IRR (95% CI) |
|---|---|---|---|
| All causes | 77 | 1005.6 | 3.1 (2.5–3.9) |
| Violence | 31 | 399.7 | 7.9 (5.4–11.3) |
| Suicide | 15 | 194 | 12.4 (6.9–20.7) |
| Non–communicable diseases | 11 | 147.6 | 1.1 (0.6–2) |
| Communicable diseases | 5 | 67.3 | 2.4 (0.8–5.7) |
| Other | 15 | 197 | 2.1 (1.2–3.5) |

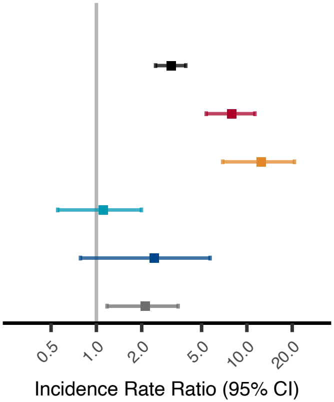


### Youth Detention

| Cause | N | Rate per 100K | IRR (95% CI) |
|---|---|---|---|
| All causes | 45 | 1079.9 | 8.1 (5.9–10.8) |
| Violence | 34 | 815.9 | 19.4 (13.3–27.5) |
| Suicide | 2 | 48 | 2.9 (0.3–10.4) |
| Other | 9 | 216 | 4 (1.8–7.6) |

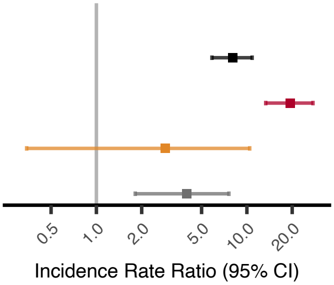
